# Prevalence and stability of SARS CoV-2 RNA on Bangladeshi banknotes

**DOI:** 10.1101/2020.11.26.20233627

**Authors:** Selina Akter, Pravas Chandra Roy, Amina Ferdaus, Habiba Ibnat, A.S.M. Rubayet Ul Alam, Shireen Nigar, Iqbal Kabir Jahid, M. Anwar Hossain

## Abstract

Originating at December 2019 in China, SARS-CoV-2 has emerged as the deadliest pandemic in the history of mankind. Along with direct contact and droplet contaminations, possibility of infections through contaminated surfaces and fomites are being investigated. In this study, we aim to assess the prevalence of SARS-CoV-2 viral RNA by real time one-step reverse transcriptase PCR on banknotes being circulating in Bangladesh. We also assessed the persistence of the virus on banknotes spiked with SARS-CoV-2 positive diluted human nasopharyngeal samples. Among the 425 banknote samples collected from different entities, 7.29% (n= 31) were tested positive for targeted genes. Twenty four representative positive samples were assessed for N gene fragments by conventional PCR and sequenced. All the samples carry viral RNA belonged to GR clade, the predominant circulating clade in Bangladesh. In the test of stability, the N gene was detected for up to 72 h on banknotes spiked with nasopharyngeal samples and CT values increases significantly with time (p<0.05). ORF1b gene was observed to be less stable specially on old banknotes and usually went beyond detectable limit within 8 to 10 h. The stability of virus RNA was well fitted by Weibull model and concave curve for new banknotes and convex curve for old banknotes have been revealed. Handling of banknotes is unavoidable; hence these findings implicated that in order to limit SARS-CoV-2 transmission through banknotes proper hygiene practice are needed.

## 1. Literature review

Severe acute respiratory virus (SARS) is a member of Betacoronavirus, a naturally circulating virus in various animals and mammals, was responsible for two previous outbreaks in just two decades; in November 2002 as SARS in China and in June 2012 as MERS (Middle East respiratory syndrome corona virus) in Saudi Arabia (Zaki et al., 2012; Zhong et al., 2003). At the end of December 2019, another SARS outbreak confirmed in China became pandemic in a short period of time. WHO named the virus as COVID-19, later renamed as SARS-CoV-2 as suggested by the International Committee on Taxonomy of Viruses (Gorbalenya et al., 2020). As on November 11, 2020, the pandemic claimed over one million death with 51.53 million confirmed cases, new areas or populations are infected daily (WHO, 2020). Several researchers have revealed that the virus mutated rapidly and new variants became predominant with different mutations for their evolutionary fitness(O. K. Islam et al., 2020)(Nextstrain:, 2020)(GISAID, 2020)(Rambaut et al., 2020). Among them, GSAID clade nomenclature has been representing the worldwide phylogenetic circulating diversity and grouped as six major clades namely GH, GR, G, V, L, and S.

Community transmission is playing significant role in SARS-CoV-2 pandemic. The direct routes of transmission includes coughing or sneezing mediated infectious droplets and mechanistic ventilation including talking or singing(Stadnytskyi et al., 2020; van Doremalen et al., 2020).Similarly, direct physical contact also is responsible for SARS-CoV-2 transmission. Another unreported and poorly understood indirect route of virus transmission is contaminated surfaces or fomites. Transmission through contaminated surface is rare, but cannot be denied. Various respiratory viruses e.g., SARS-CoV, Influenza, Rhinovirus etc. can persist in various fomites without losing infectivity for a certain period (Thomas et al., 2008). After the SARS-CoV-2 pandemic, researchers are reporting presence of SARS-CoV-2 RNA on various surfaces such as patients’ masks (Li et al., 2020); door handles and sanitizer dispenser (Razzini et al., 2020); different surface areas of hospital (Santarpia et al., 2020); sewage pools (Wang et al., 2020). Unlike previous pandemic causing coronaviruses (e.g., SARS and MERS), SARS-CoV-2 has higher reproductive number (*R*o), an indicator of virus transmissibility, thus is highly contagious in nature and has low incubation period but higher infection rate (Xie and Chen, 2020). To control the outbreak, it is pertinent to identify the potential source of infection in community, characterize the route and break the chain of transmission. Like other fomites, banknote is an ideal source of pathogenic microbes (Gabriel et al., 2013; Hiko et al., 2016; Jalali et al., 2015; Maritz et al., 2017; Pal and Bhadada, 2020; Thomas et al., 2008).Microbes remain infective in papers, stability depends on initial loads, temperature, and moisture content of the surrounding environment (Pastorino et al., 2020). Banknote is the most ubiquitous and transferable object in present world. Countries like Bangladesh have limited user of virtual banking and are dependent on paper banknote. Although few countries decontaminate their paper notes in regular interval and assess the microbial contamination, which is seemingly not possible for many nations. The reasons behind microbial contamination in paper notes are material composition specifically moisture absorbing and dust retaining capacity and high frequency of exchange (Vriesekoop et al., 2010).The rough surface of paper based banknote also provides the necessary support for microbes to settle down and be accumulated. A recent report showed that SARS-COV-2 survives on banknotes for 28 days at 20 °C (Riddell et al., 2020). However, best of our knowledge, there is no data regarding the prevalence of SARS CoV-2 on banknotes as it can survive longer (Riddell et al., 2020). Although microbial survival on different surfaces as linear regression (Riddell et al., 2020), but many researchers documented the microbial survival as nonlinear, concave or convex with sigmoid shapes (Buzrul and Alpas, 2007; Coroller et al., 2006; Jahid et al., 2013; Mafart et al., 2002). In this research, we aimed to find out the prevalence of SARS-CoV-2 RNA on circulating Bangladeshi banknote and to assess the stability of SARS-CoV-2 on spiked banknotes.

## 2 Materials and Method

### 2.1 Ethics approval

The work has been conducted in the Genome Centre of Jashore University of Science and Technology, which is providing SARS-CoV-2 real time RT PCR diagnostic service as part of national COVID 19 response and surveillance program (https://dghs.gov.bd/images/docs/Notice/rt_pcr_lab.pdf). The research project has been reviewed by the institutional ethical review board of Jashore University of Science and Technology with an exemption of consent from the patients (ref: ERC/FBS/JUST/2020-42).The non-issuable banknotes used in this study were collected from a public bank, which were withdrawn from the circulation for destruction.

### 2.2 Sample Collection and processing

We have collected circulating banknotes of varying denominations as an exchange of payment from retail shops, ticket vendors and auto rickshaw drivers etc at places in two Southern districts namely Khulna and Jashore, of Bangladesh. During collection of banknotes, we avoid selection bias. Hence, the banknotes are not equaly distributed but representing the real circulation frequencies among denominations. Sample collector asked to put the paper banknotes directly in a biohazard sample collection bag and zipped the bag instantly. Samples were transported instantly to the laboratory, preferably within 2 hours (h). Each of the banknotes was removed from the bag in a biosafety Class IIa cabinet and kept on sterile polystyrene petri plate. Both the sides of the banknote was swabed with a nylon flocked round (oral) swab (Biovencer Healthcare Pvt. ltd., India) soaked with 0.9% saline containing Rnasin (Sunsure Biotech, China). The head of the swab is cut down and put into 80.0 μl of 0.9% saline containing Rnasin in a Dnase/Rnase free microcentrifuge tube and mixed vigorously. Twenty microliter of the resuspended sample was aliquotted into a new microcentrifuge tube, 20.0 μl of ice cold RNA extraction buffer (QuickExtract™ RNA Extraction Kit, Lucigen) was added to it and vortexed for 1 minute. The extract was immediately chilled on ice and store at -20°C till real time RT PCR amplification of SARS-CoV-2 specific genes.

### 2.3 Preparation of banknote notes spiked with SARS CoV2 positive sample

#### 2.3.1 Experimental set up

To assess the stability of SARS-CoV-2 RNA on paper banknote, we have selected three SARS-CoV-2 positive human samples from Genome Centre of Jashore University of Science and Technology containing both nasal and oro-pharyngeal swab in 0.9% saline water. The Center is routinely diagnosing the COVID-19 patients’ samples by real time RT PCR method (Sansure Biotech, China). All the three samples were selected among SARS-CoV-2 positive (CT= 22)routine diagnosis samples. We used six different banknotes, three were visibly old and dirty circulated for years and the other three were new and clean but tore apart. Banknotes were collected in zipped locked bag, cut into circular pieces of 28sq mm (3mm of radius) size and three random pieces from each banknote were tested for SARS-CoV-2 RNA by real time RT PCR method and found negative. All the pieces of banknotes were placed in polystyrene Petri dish and spiked with 10 μl of two-times diluted SARS-CoV-2 positive samples (39 pieces of each banknote types were spiked with each of the three types of positive samples), allowed to dry in a biosafety IIa cabinet. When the banknotes were visibly dry, the Petri dishes were placed inside a clear biohazard bag and sealed. All experimental sets of banknoteswere kept inside the biosafety IIa cabinetat room temperature with negative pressure. After setting the whole experiment, three replica spiked banknote pieces were picked from each set and kept in micro-centrifuge tubes at different time intervals (i.e., 0h,1 h, 2h, 4h, 6h, 8h, 10h, 12h, 24h, 36h, 48h, 60h and 72h). Temperature was monitored and recorded throughout the experiment by room thermometer (Model: SHX-RPT-6, Shanghai, China). Suitable biosafety precautions (e.g., overhead gown with N95 respirator) were taken by the researcher throughout the work and suspected samples as well as spiked banknotes were handled only in BSC IIa facilities.

#### 2.3.2 RNA extraction from spiked banknote

Twenty microliter of 0.9% saline was poured in microcentrifuge tube containing each banknote piece, vortex mixed and kept on ice. Twenty microliter of ice cold RNA extraction buffer (QuickExtract™ RNA Extraction Kit, Lucigen) was added to it and vortexed for 1 minute. The tube was immediately chilled on ice and stored at -20°C. Before perforrming real time RT PCR amplification of SARS-CoV-2 specific genes, the tubes were thawed, spined shortly and aspirate 10 μl carefully to be used as template.

### 2.4 Detection of SARS-CoV-2 RNA

We used the primer and probe sequences and protocol designed and used by Chu et al.(2020). The sequence of primers and probes used for this study has been listed in Table A1. Due to the lack of suitable positive control, RNA extracted from a previously characterized (whole genome sequenced, accession no.EPI_ISL_561377) positive sample was used as positive control and 0.9% saline prepared with RNase free water was used as negative control in this assay. The GoTaq® Probe 1-Step RT-qPCR Reaction Mix (Promega, USA) was used for the duplex reaction and the reaction mixture was prepared following protocol provided with the kit, with an exception of using 9 μl RNA extract as template and preparing final reaction volume of 25 μl (instead of 20μl). The thermal Cycling parameters were optimized as followed: Reverse transcription at 45°C for 30 minutes; Reverse transcriptase inactivation and GoTaq® DNA Polymerase activation at 95°C for 2 minutes; and 45 cycles of the regime of denaturation at 95°C for 15 seconds and Annealing with extension at 60°C for 1 minute. The PCR reaction was performed in a QuantStudio™ 3 Real-Time PCR System (The Applied Biosystems, USA) in 96-well plate (0.2 mL). The result was analyzed in Quantstudio design and analysis software(v1.3.3). Sigmoid curve for either or both of the *orf1b-nsp14* or *n* gene with a CT value of ≤ 36 was interpreted Positive. CT value between 36 and 39 were repeated and above those were considered negative in the prevalence study. For the assay of stability of SARS-CoV-2 RNA on banknotes, the half-life of the viral RNA was measured in different experimental set up by comparing with standard curve created by a serial of 2-fold dilutions of the inoculum by the identical RNA extraction and real time RT PCR protocol.

### 2.5 Preparation of cDNA for targeted PCR

From the leftover RNA, we prepared cDNA for the representative 21 samples using the ProtoScript® II First Strand cDNA Synthesis Kit (NEB, UK). We omitted the ‘denaturation of RNA secondary structure’ step wherein we mixed 6µl of extracted RNA with 2µl of random primers and other components are used as mentioned in the protocol. Then, the mixture was annealed and incubated at 42°C and 48°C for 5 and 20 minutes, respectively, followed by deactivating enzyme at 80°C for 5 minutes and immediate chilling on ice. The final reaction mix was 20μl for each cDNA synthesis reaction to be performed.

### 2.6 Determining the intactness of viral RNA genome on bank notes

For checking the intactness and possibly the infectivity of the virus, we targeted an 850bplarge segment of the viral genome, which spans the receptor binding region (RBD) of spike protein coding sequence. The forward and reverse primers are S_F2 (GCTGTAGACTGTGCACTTGACCC) and S_R2 (GTAGTGTCAGCAATGTCTCTGCC), respectively. We carried out the PCR in 10µl reaction volume comprising of 4 µl cDNA, 5µl hot-start color master mixture (GoTaq® G2 Green Master; Promega, USA), 0.5 µM of each forward and reverse primer. The thermocycling conditions were maintained as the initial denaturation at 95°C for 1 min followed by 35 cycles at 95°C for 30s, annealing at 56°C for 30s, and 72°C for 50s followed by a final extension at 72°C for 5min. Finally, we electrophoresed PCR products on an 1% (w/v) agarose gel stained with ethidium bromide (UltraPure™ Ethidium Bromide, 10 mg/mL; Thermo Fisher, USA) and visualized using a gel documentation system (Bio-Rad, USA).

### 2.7 Targeted (Sanger) sequencing of SARS-CoV-2 gene

The randomly amplified cDNAs were used as template for the PCR targeting N protein coding sequence for detecting the viral phylogenetic clade (M. T. Islam et al., 2020). After purifying the PCR products with the ExoSAP-IT™ PCR Product Cleanup Reagent (Thermofisher Scientific, USA) as per manufacturer’s instruction, the BigDyeTerminator v3.1cycle sequencing ready reaction kit (ThermoFisher Scientific) was used in a way to optimize the cost than as mentioned in the Islam et al. (2020). Instead of 0.5 μL, 0.25μL (per 10μL reaction) undiluted BigDye Terminator v3.1 Ready Reaction mix was used together with 1 μL 5X sequencing buffer, 0.3 μL primer, 3μL template DNA and 5.7 μL nuclease free water. We setup the cycle sequencing PCR condition according to the kit protocol. We performed the further bioinformatics analyses considering Wuhan-Hu-1 (NC_045512.2) as the reference sequence using Molecular Evolutionary Genetics Analysis (MEGA X) software (Kumar et al., 2018).

### 2.8 Scanning Electron Micrograph

Representative disc of new and old banknotes were air dried, mounted on SEM sample loading disc with carbon tape and sputter coated with gold using the timed gold sputter preset recipe in Quorum Q150RS plus machine at following conditions: sputter current of 20mA, sputter time of 120s with tooling factor 2.30. The micrograph was taken in Gemini Sigma 300 field emission scanning electron microscope and digitized images were collected by Smart SEM software (Zeiss, Germany).

### 2.9 Non-linear regression

Generally Weibull model used to estimate different parameters which are non-linear model. The equation is following:

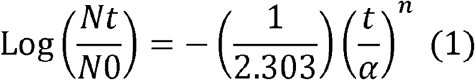

Where α is called scale parameters (unit is min or sec) and *n* is shape parameters (unit less) (van Boekel, 2002), *Nt* number of microorganisms (CFU/mL or cm^2^) after survival time *t, N0* initial number of microorganisms, *t* is the survival time of microorganisms (hr). *n* values equal to 1 correspond to linear survival curves, *n* values > 1 corresponds to downward concave survival curves, *n* values < 1 corresponds to convex survival curves.

However, equation (1) can be reparametrized by Coroller et al., (2006) as the following equation:

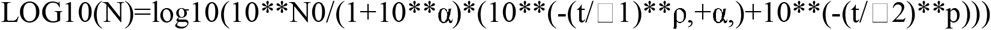

Here,α = log_10_(*N*_01_/*N*_02_)is the difference of two subpopulation, □1 is the scale parameters of first subpopulation,ρ is the shape parameter and δ2 is the scale parameter of second subpopulation (Coroller et al., 2006). For the old banknotes, the equation (1) can be reparametrized by Mafart et al., (2002)as follows:

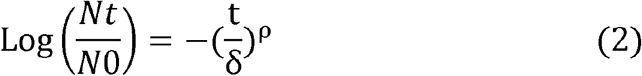

Here, ρ is the shape parameter and δ is the scale parameter (Mafart et al., 2002).

If the model fits properly,a linear shape if *p* is equal to 1, a concave shape if *p* is >1, and a convex shape if *p* is <1. Root Mean Square Error (RMSE) was calculated from the software to know the goodness of fit of the model to death rate. The RMSE values closer to 0 and adjusted R^2^ values close to 1.0 indicates better fitness of model. The model has been for new banknotes.

### 2.10 Statistical analysis

To determine the prevalence and stability of SARS-CoV-2 RNA on banknotes, all experiments were repeated three times. CT values from the stability on banknotes have been transformed as 1/CT*100 and were plotted with respective time (hr) by using Microsoft Excel 2010Add-in GInaFiT 1.6 (Geeraerd et al., 2005) (https://cit.kuleuven.be/biotec/software/GinaFit).

The α, □1, ρ, □2 values were analyzed by carrying out ANOVA by using SAS software version 9.2 (SAS Institute Inc., Cary, NC, USA) for a completely randomized design. When the effect was significant (p< 0.05), separation of the mean was accomplished with Duncan’s multiple-range test.

## 3. Result and Discussion

### 3.1 Prevalence of SARS-CoV2 on banknotes

A total of 425 (n) banknotes were collected from 56 (N) entities over a period of three months. The entity includes Pharmacies, ticket vendor/collector and drivers of local and inter-city transports, various shops and restaurants (Table 1). Among the entity, we have collected 7.58 banknotes on an average (minimum 2 and maximum 19 banknotes per entity, data not shown). In total, we found 7.29% (31/425) banknote samples were positive for SARS-CoV-2 RNA assessed by real time one-step RT PCR method. The entity of local transport (N=11, n=84) includes banknotes from drivers of three wheelers(e.g., auto rickshaw, and mechanically driven three-wheeler), which accounted for the highest prevalence (14.28%) among the entities. In a case, seven SARS-CoV-2 RNA positive banknotes (out of 19) were found from a particular auto rickshaw driver, which contributed to the overall higher prevalence rates among the banknotes collected from local transports. On the contrary, banknotes sampled from the ticket vendors and - collectors at inter-city transport (bus) were negative for the viral RNA. During the study, intercity transport authority were regulated to ensure wearing masks, maintain social distancing (carrying 50% of total capacity) with personal hygiene.Prevalence of SARS-CoV-2 RNA was detected on around 8 to 10 per cent ofbanknote samples collected from restaurants& food shops (N = 5, n=38) and grocery shops (N=13, n=106). The detail result has been listed in Table 1. The study was designed and started at time when the number of SARS-CoV-2 infected confirmed cases was at the peak (end of June 2020) in the country. The weekly cases of new SARS-CoV-2 infection was around 21K at July and declined periodically to around 5K at the end of September, 2020 (Figure A1).

**Figure 1.**
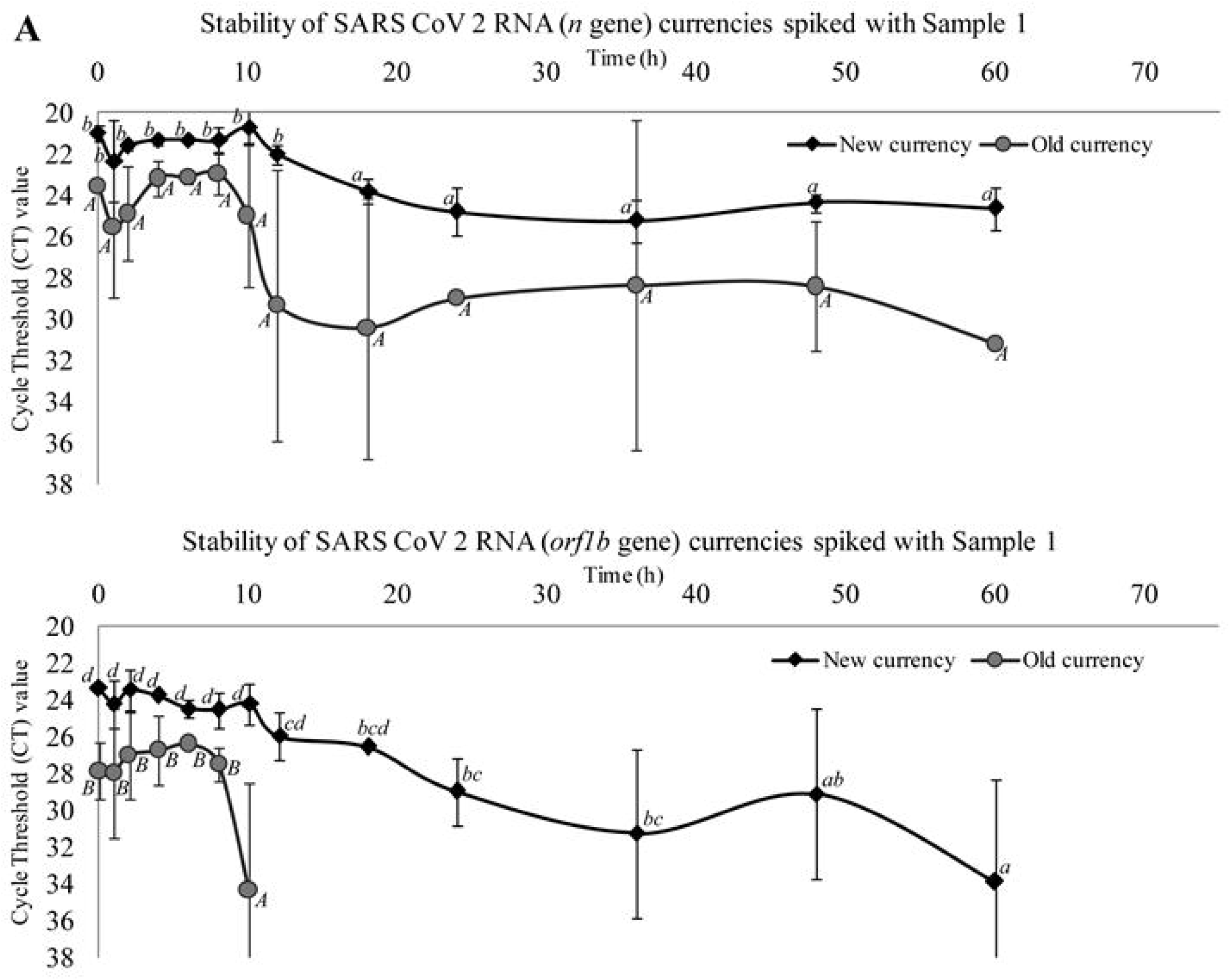
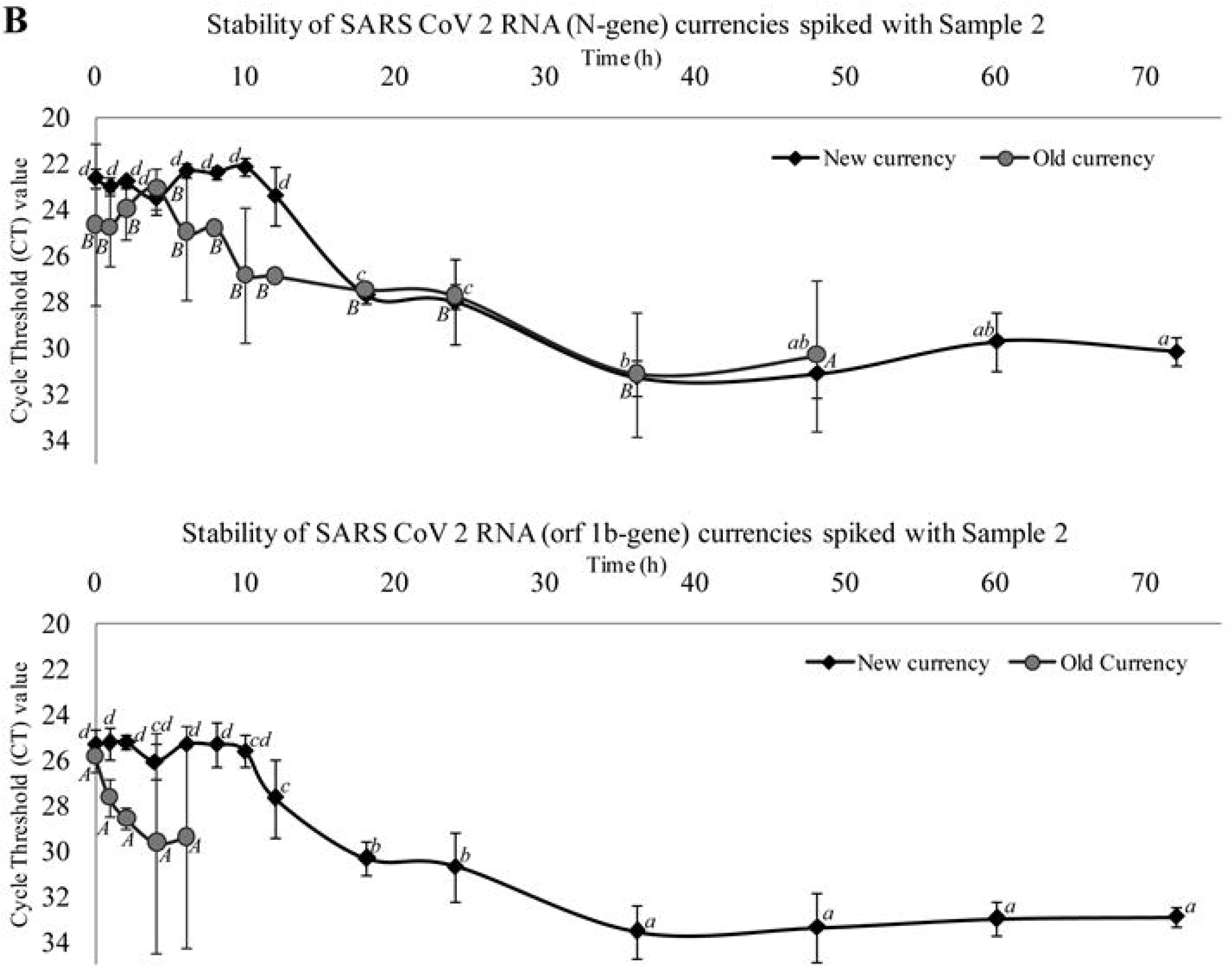
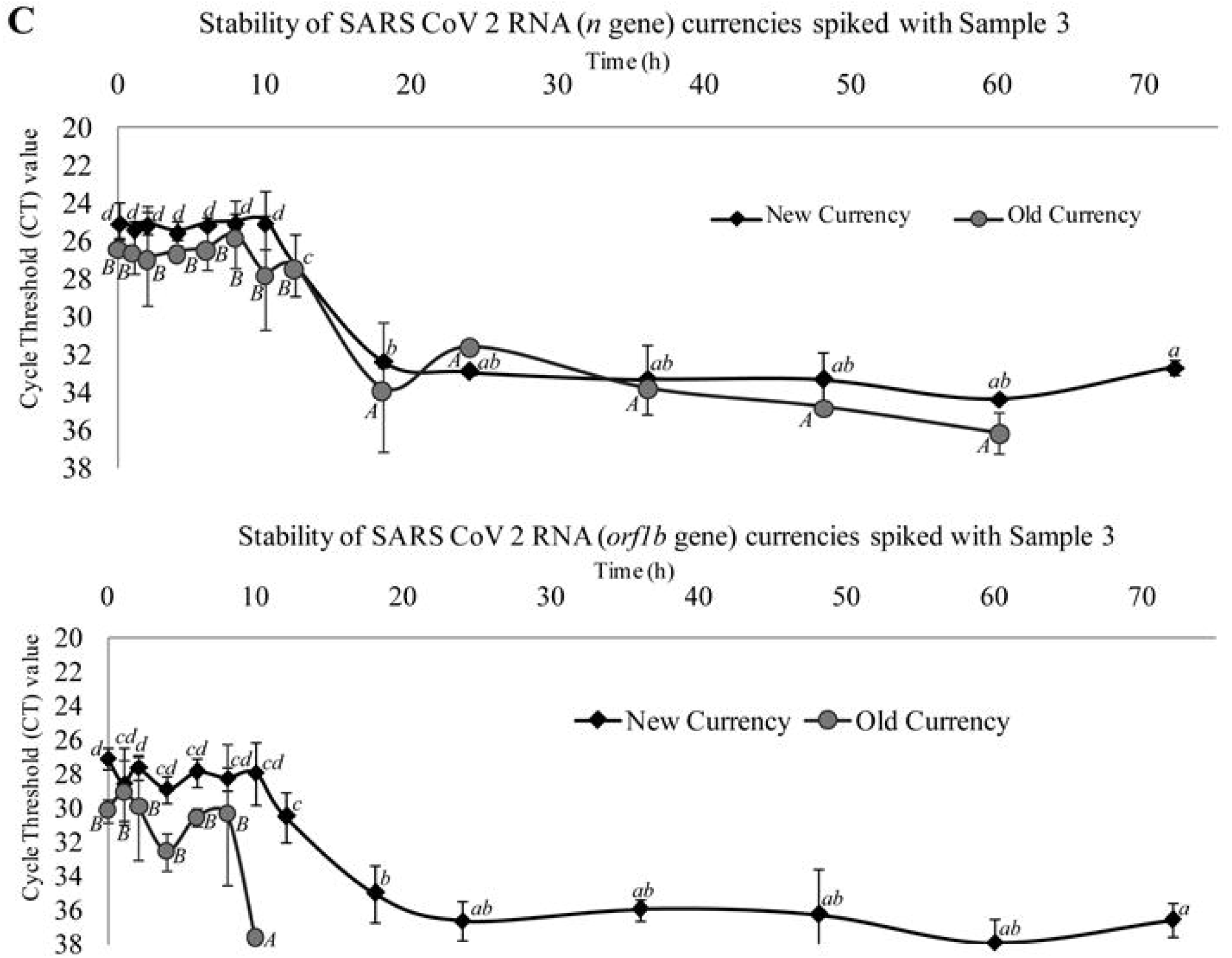
Stability of SARS-CoV-2 on currencies spiked with crude samples: Line graphs representing the changes in CT values of ‘n’ and ‘orf1b’genes on new and old currencies (non-issuable) spiked with Sample 1 (fig. A), 2 (fig. B) and 3 (fig. 3) at different time intervals (staring from 0h to 72h). Values are the mean (with standard deviation as error bars) of three independent replica experiments. Within each variable, values with the same letter are not significantly different according to Duncan’s multiple range test (P. 0.05); lower case letters used for values of experiments on new currencies and upper case letters for values of experiments on old currencies.

**Table 1:** Prevalence of SARS CoV-2 RNA on circulating Bangladeshi currencies collected from different entities.

| Category/<br>Name of<br>Entity | Number<br>of Entity<br>(N) | Number of<br>Positive<br>Entity | Number of<br>currency (n) in<br>each category | Number of<br>positive in each<br>category | Category<br>wise<br>positive % |
| --- | --- | --- | --- | --- | --- |
| Pharmacy | 7 | 1 | 67 | 2 | 2.98 |
| Local<br>Transport | 11 | 4 | 84 | 12 | 14.28 |
| Intercity<br>Bus | 6 | 0 | 41 | 0 | 0.00 |
| Others shop | 14 | 2 | 89 | 2 | 2.25 |
| Grocery<br>Shop | 13 | 5 | 106 | 9 | 8.49 |
| Hotel/rest. | 5 | 3 | 38 | 5 | 13.15 |
| Total: | 56 | 15 | 425 | 31 | 7.29 |

In this study, we collected banknote in a random basis which included higher denomination (1000 TK, 500 TK, 100 TK) (n= 22), medium denomination (50 TK and 20 TK) (n = 143), and lower denomination (10 TK, 5 TK, 2 TK) (n = 260) banknotes. Although the sample distribution was not uniform, but we found higher prevalence in higher denomination (13.63%, 3/22) compared to medium (4.89%; 7/143), and lower denomination banknotes (8.07%; 21/260). The higher denomination banknotes usually have less transaction frequencies and cleaner than middle or lower denominations, the higher prevalence in this group were not predicted but could be due to the larger surface area. However, we found that new clean banknote supports more stability or recovery of SARS-CoV-2 RNA than the older one (discussed later). In terms of monthly distribution, the samples were not similar but the positive cases per cent samples were apparently homogenous (Table 2). However, the samples were collected only for three months; hence Levene’s test could not be performed to calculate the homogeneity of variance.

**Table 2:**
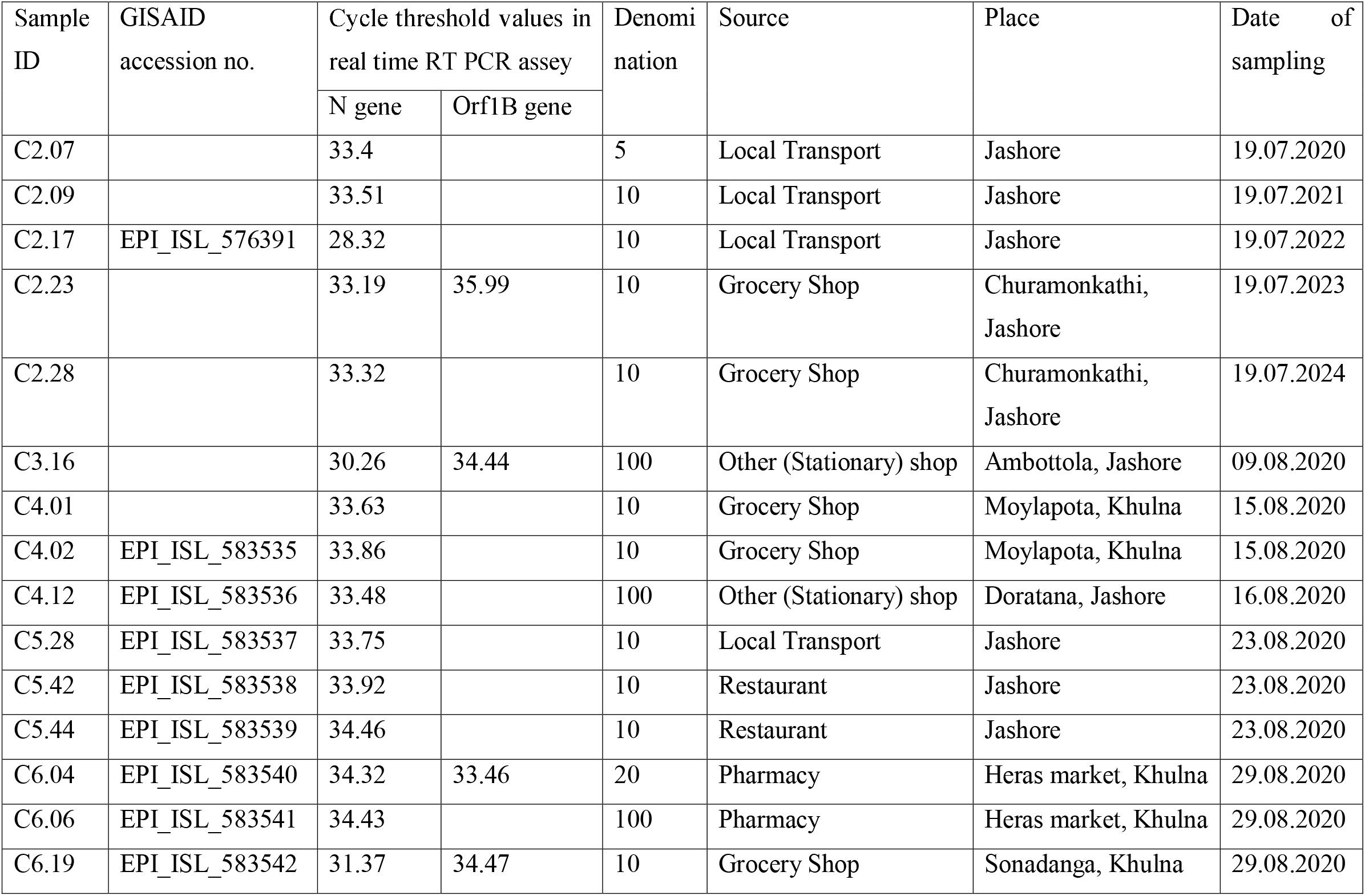
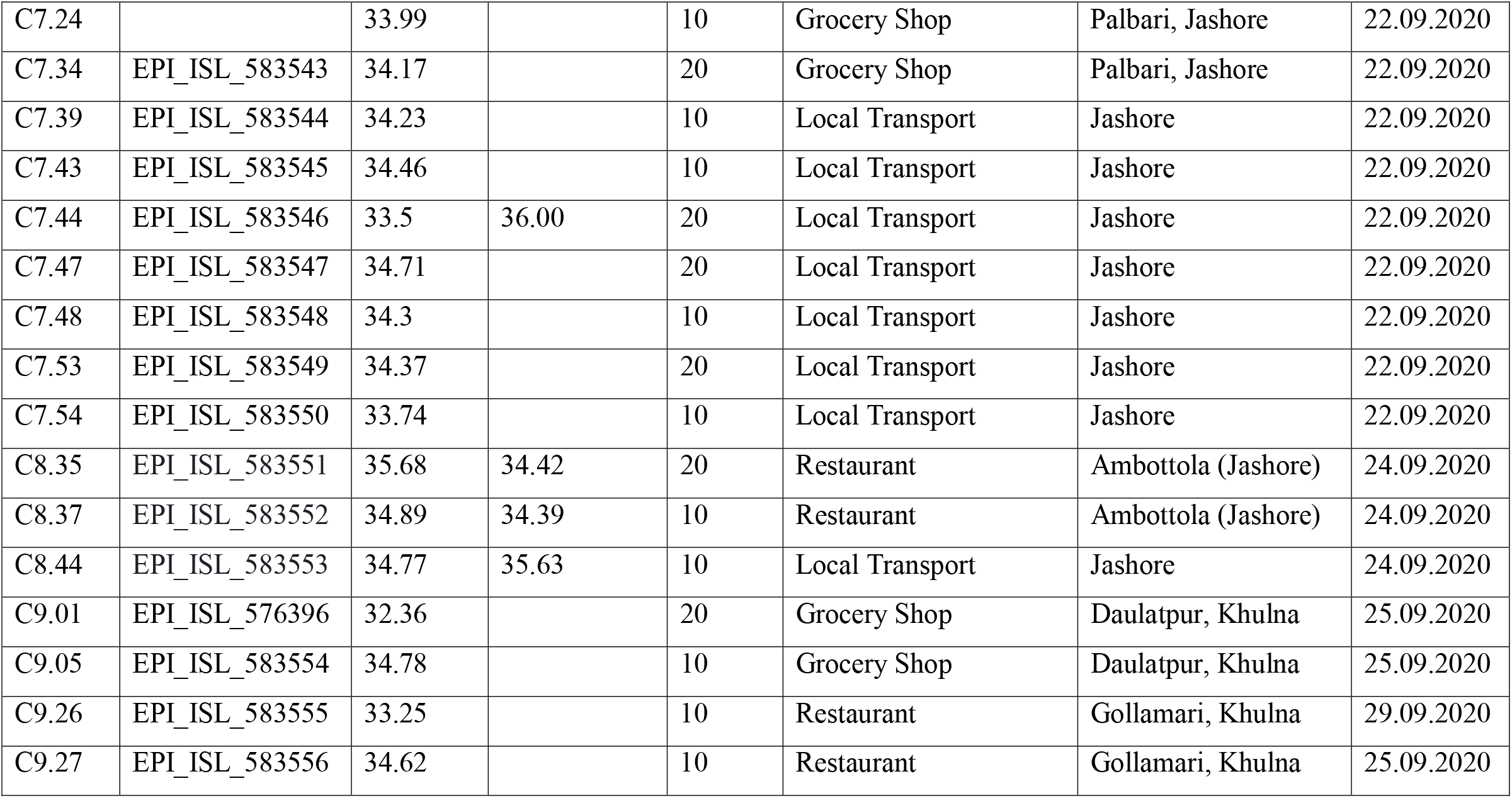
SARS-CoV-2 positive (detected by Real time RT PCR) currency distribution

All the samples were collected from the environment not related to hospital or isolation center for SARS-CoV-2 patients. The presence of SARS-CoV-2 RNA on the circulating banknotes indicates the frequent movement of infected patients either asymptomatic or with mild symptoms. Milder symptomatic patients may have higher viral load (low CT value) and can shed virus(Jan et al., 2020). In a recent survey on2157 human subjects in Bangladesh found out that 16.3% of the people do not wear mask and 24.6% do not avoid crowd, even having milder SARS-CoV-2like symptoms (Hossain et al., 2020). Detection of viral RNA does not ensure presence of infective viral particle but at the same time does not exclude the possibility of having it.

### 3. 2 Gene seuqncing

Randomely selected banknote samples, tested positive for SARS-CoV-2 RNA in real time RT PCR method, were also amplified for different segments of other genes by conventional PCR method and PCR products of 24 representative samples were sequenced. Sequence data evaluated that all the samples contained SARS-CoV-2 virus belong to GR clade strains.The result signifies the dominant presence of this clade in Bangladesh as described in other studies (Alam et al., 2020). In Bangladesh, 73% (273/372) of the viral strains are of GR clade as per GISAID sequence information whereas 38% (58,969/1,56,381) of the viruses are of the clade worldwide. Accession IDs to the submitted sequences as an archetype are available in GISAID EpiFlu™ database (Table 2). Noteworthy, we did not find any long amplified products, i.e. 850 bp targeted amplicon spanning RBD region, for each of the samples (data not shown) that states the lacking of intactness of the viral genome on bank note, thus infectivity.

### 3.3 Stability kinetics of spiked samples

The survival kinetics of spiked samples of present study on new and old banknotes were determined using Microsoft Excel 2010 Add-in GInaFiT 1.6 (Geeraerd et al., 2005) (https://cit.kuleuven.be/biotec/software/GinaFit). The graphs show the survival populations of SARS CoV-2 with respect to time (h) of N gene for new banknotes (Fig 2A), N-gene for old banknotes (Fig 2B), orf gene for new banknotes (Fig 2C) and orf gene for old banknotes (Fig 2D). The results revealed that overall survival of SARS-CoV-2 on new banknotes were higher compared to older banknotes. The N gene stability was higher compared to orf gene. For N gene in new banknotes, for the all three samples survived up to 60 h but absent after 72 h (Fig 2A).

**Figure 2:**
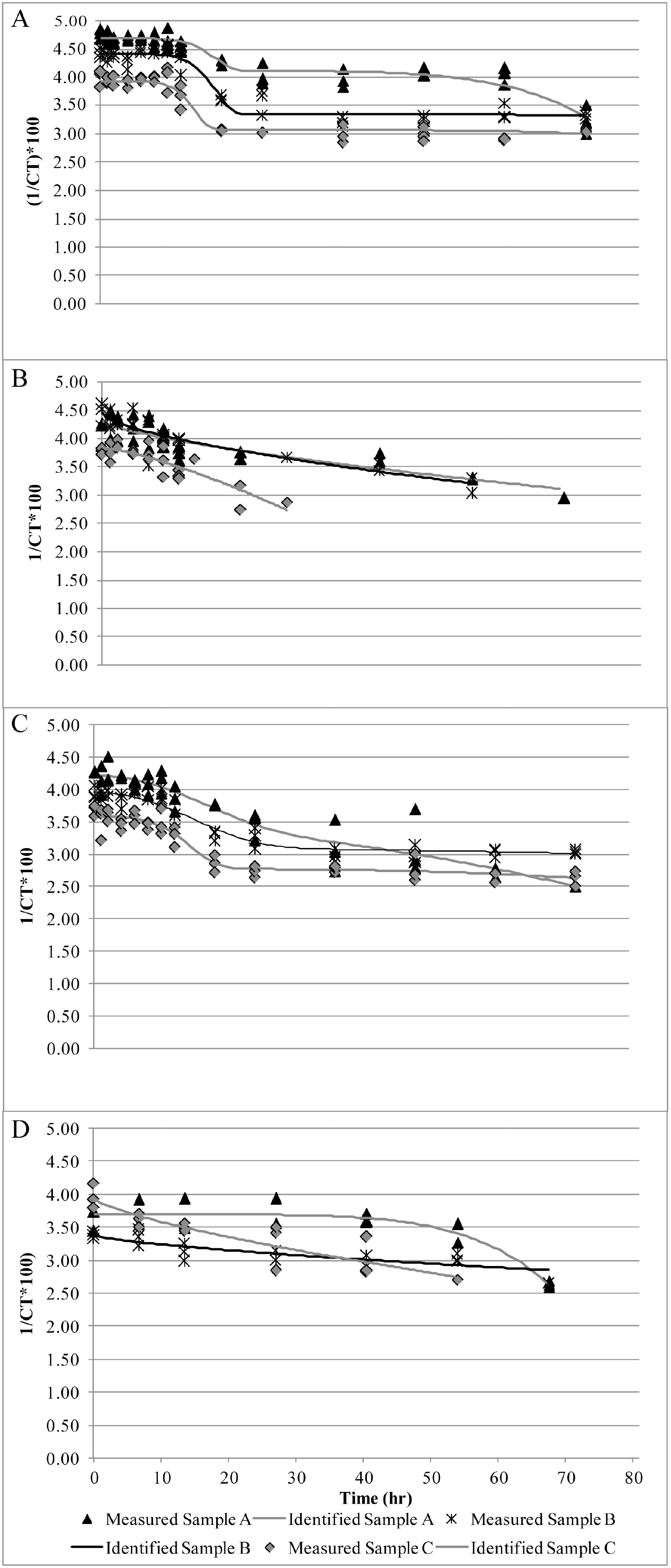
SARS-CoV-2 Stability on spiked diluted nasopharyngeal swab on new and old banknotes and incubated at room temperature and samples were tested by RT PCR at different time. The points represent mean ± SEM of 3 independent experiments, and vertical bars indicate the duration of time to keep the samples. Curves were fit to a modified Weibull model using GInaFiT 1.6 software. (A) curves for *n* gene of new banknotes of samples A, B and C, (B) curves for *orf1ab* gene of new banknotes of samples A, B and C (C) curves for *n* gene of old banknotes of samples A, B and C (D) curves for *orf1ab* gene of old banknotes of samples A, B and C

The results also revealed that for new banknotes, the values did not decrease upto 10 h and then gradually decrease up to 60 h. For the old banknotes, the n gene stabilty were less and at 60 h the values were zero (Fig 2B). For old banknotes, except one samples, other two samples did not show any stability for initial 10 h and later hours. The graph shows a sharp decrease in case of old banknotes (Fig 2B). All the samples were stable upto 60 h for orf gene (Fig 2C) whereas only 10 h for old banknotes (Fig 2D). In new banknotes, the graph show the concave while old banknotes graphs show the convex type. Therefore, it has been hyphothesized that SARS COV-2 in old banknotes are less stable. Our results of stability of SARS-CoV-2 on new banknotes shows similarity with the finding of Kampf et al., (2020). The researchers found highest 5 days stability for SARS-CoV on paper surfaces. Riddell et al., (2020) found that the virus survived less than 7 days on bank notes at 30°C and one day at 40°C. Our laboratory temperature was around 35°C during the experimental conditions. Even though we used the samples from patient’s nasopharyngeal swab whereas the authors used the virus grown in Vero cell line, however, the results of our study agreed with Riddell et al., (2020) and Kampf et al., (2020). SARS-CoV-2 has been reported to stay infective in various inanimate objects (e.g., metal, plastics etc.) for 2 hours to 9 days (Kampf et al., 2020). In another study, coronavirus exposed to various metals (Copper or copper alloy) have negative impact on virus, irreversible damaged intactness of virus and RNA (Warnes et al., 2015). Temperature and relative humidity can negatively affect the stability of SARS CoV-2 and reduce the transmissibility over time (Demongeot et al., 2020) which may be true for other respiratory enveloped virus, but this may be still inconclusive (Ma et al., 2020; To et al., 2021), more research needs to be performed for the SARS-CoV-2.

### 3.4 Mixed Weibull kinetics of SARS-CoV-2 spiked on bank notes

Table 3 shows the Weibull model parameters and goodness-of-fit values of news and old bank notes spiked with SARS-CoV-2. It has been revealed that the Weibull model accurately predicted with both N gene and ORF 1b gene, with adjusted correlation coefficients (R^2^) of ≥0.8.0. Estimated RMSE was <0.3, meaning that the Weibull model was a good fit for all the survival curves for both new and old bank notes. As in the Weibull model, α,δ1, *p* andδ2 influence the curves and fitness of the data. The analysis shows that α parameters were significantly different (p ≤ 0.05) according to ANOVA which means that subpopulations 1 and 2 for new were significantly different. We found that for samples 1, 2 and 3 for n gene and orf gene of new bank notes, differences in α, δ1, δ1, *p* and δ2 were not significant (p > 0.05). The same parameters were non-significant (p≥0.05) for old bank notes. Most of the cases of old bank notes, the *p* values were □1.0 mean the convex curve for the old notes (Table 3). As the virus was most unstable, therefore, for old notes the curves were fitted by Mafart et al., (2002)not by double Weibull of Coroller et al., (2006).Suman et al., (2020) reviewed the stability of SARS-CoV-2 on different surfaces and none of the surfaces shows more than 72 hr survival time. Even the authors demonstrated that infection capability are linear, however, their graphs shows initial stability and then gradual decrease of the virus. Kwon et al., (2020)analyzed the virus on different surfaces and found the linear reduction of the virus. Riddell et al., (2020)also analyzed the survival curve with linear regression but at 20°C the graphs seem non-linear.

**Table 3.**
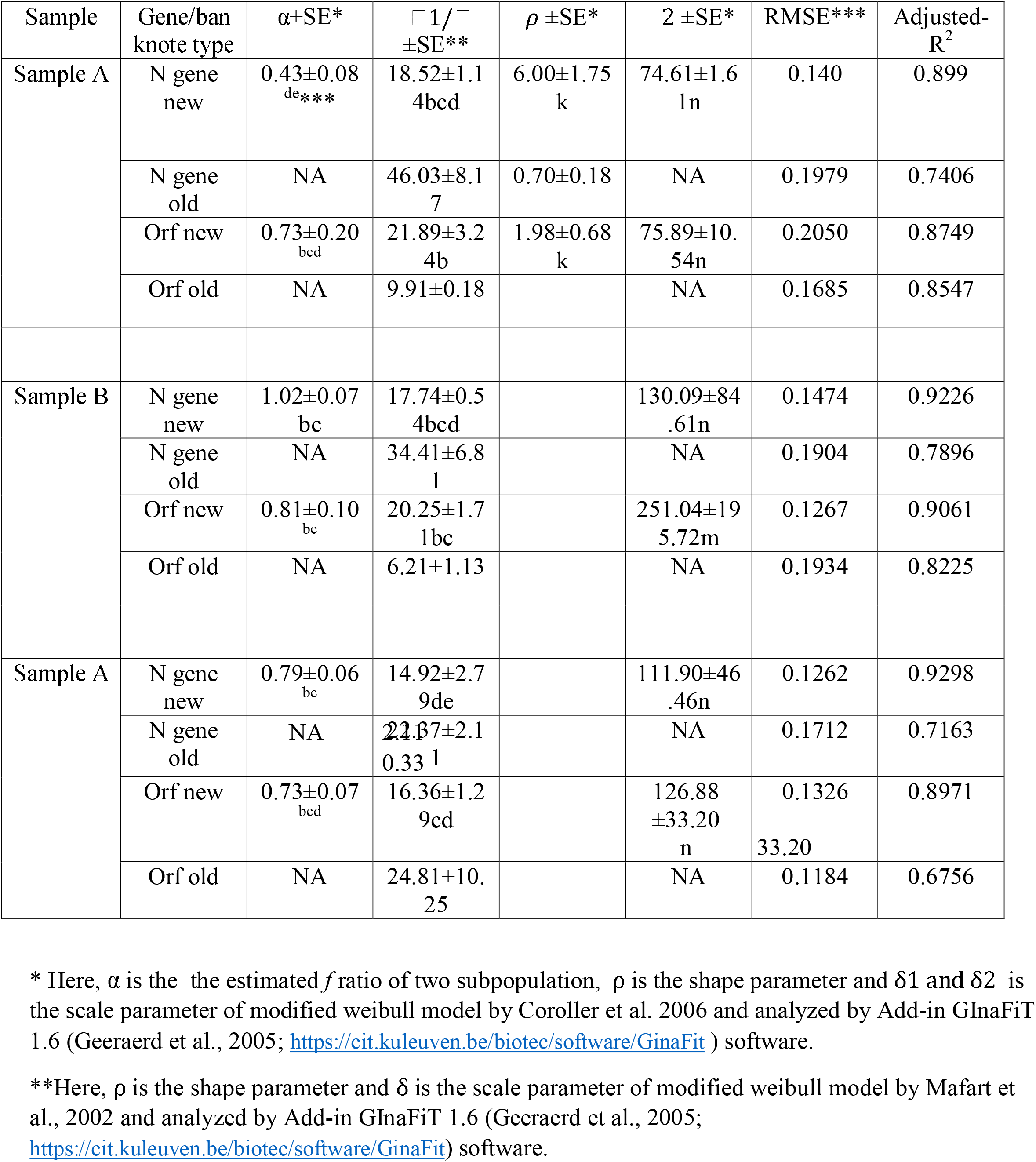
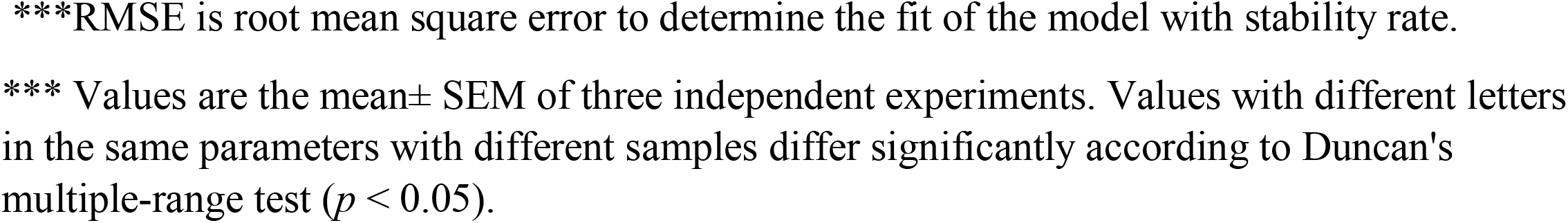
Weibull model parameters for spiked bank notes with SARS-CoV-2

### 3.5 FESEM

The observation stipulates the possibility for the presence of exogenous RNAses sourced from perpiration or dirts on banknotes. We could not assess the infectivity of the contaminating viruses, however the dirtier and old banknotes are less likely to support the stability of SARS CoV 2 RNA. The scanning electrom micrograph (Fig. 3) revealed that the old banknotes (prior to spike with SARS-CoV-2 positive human sample) are almost covered by loose layer of dirt, which possibly provide rooms for the predator and harbor for the exogenous RNAses. on the other hand, the new banknotes were observed to have more fibrous and compact texture and seemed to be more absorbant.

**Figure 3:**
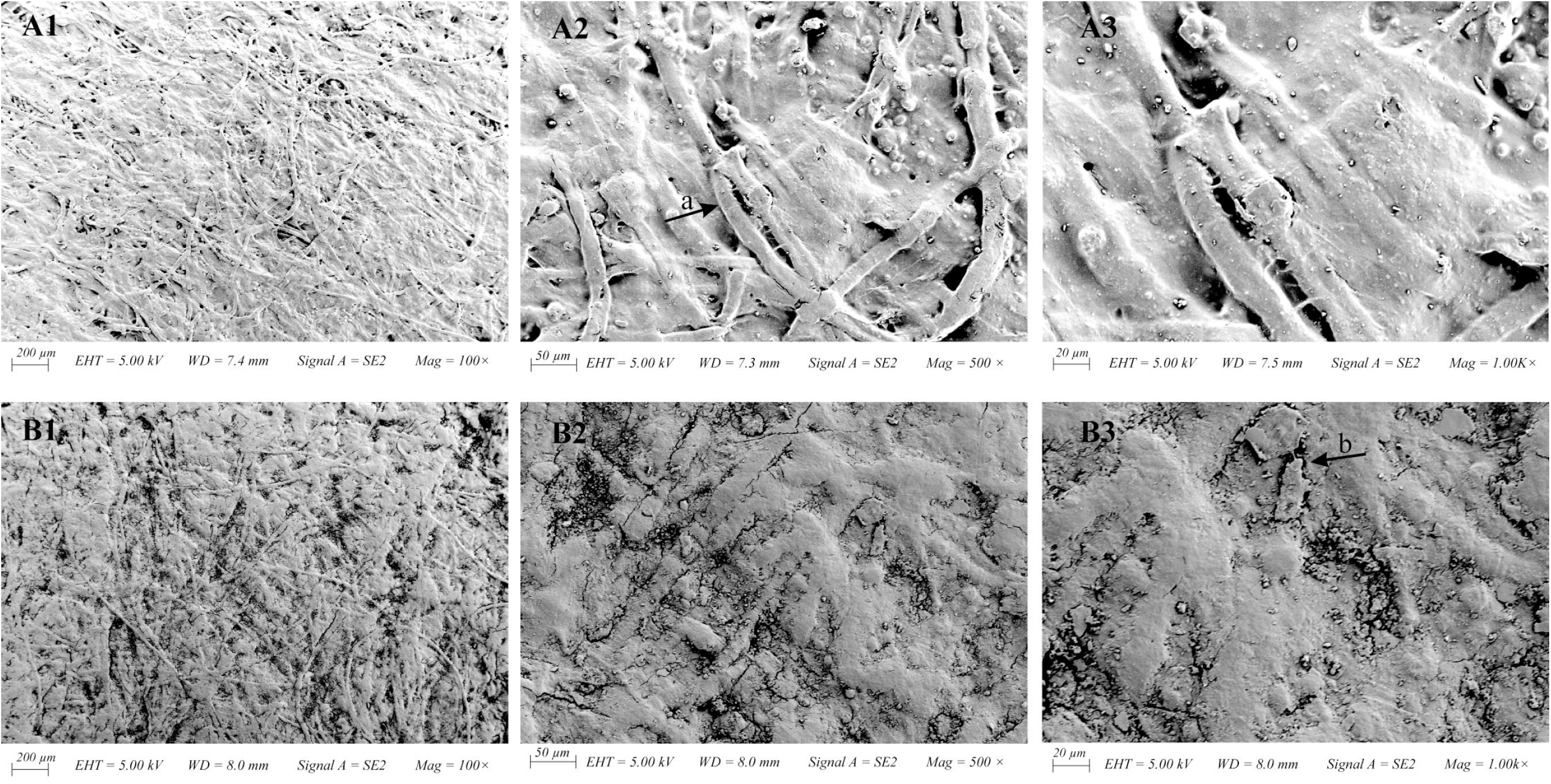
Field Emission Scanning Electron Micrograph of New (A1 to A3) and Old (B1 to B3) Bangladeshi Banknotes (representative fields) at different magnifications. Emission and scanning parameters are shown at bottom of each field in the figure. The arrow-a has pointed to show the fibrous structure of the paper banknote, and arrow-b points loosely attached dirt particles on the banknote.

## 4 Conclusions

The presences of SARS CoV-2 RNA in various environmental samples and surfaces have profound effect on viral epidemiology and infection. In our research, we found a significant portion of Bangladeshi banknotes is contaminated with either virus or RNA. Fomites like banknote mediated transmission could exaggerate the overall situation, and posed risk for the susceptible individuals. Appropriate maintenance of personal hygiene, avoidance of touching unnecessary surfaces, washing hand after handling banknotes and an effective decontamination strategy might prevent the fomite mediated SARS-CoV-2 community transmission.

## Data Availability

not available

## 5. Acknowledgements

The research has been conducted under the fund allocated from Jashore University of Science and Technology through University Grants Commission (UGC) of Bangladesh. Special thanks to Directorate General of Health Services, Ministry of Health & Family Welfare, Bangladesh for approving Genome Centre of JUST to provide real time RT PCR diagnostic service in the national COVID 19 response strategies. Thanks to Sonali Bank Limited, Corporate Branch, Jashore for providing non-issuable banknotes to support the research.

## 6. Author contributions

SA conceived the idea, performed the experiment, analyzed the data and help to write draft manuscript; PCR performed the experiment, analyzed the data and wrote the draft manuscript; AF collected the entire samples and with the help of SN extracted RNA for RT PCR assay; HI and ARA performed the experiment involved in PCR, operation, analysis and deposition of the gene sequences; IKJ and MAH contributed significantly to the research design, supervision, result interpretation, statistical analyses and improvement of the manuscrpit to its finished version.

