## Supplementary material for "Prevalence and stability of SARS CoV-2 RNA on Bangladeshi banknotes": Supplementary material.docx

APPENDICES

Table A1: List of primers and probe sequences used for the real time RT PCR detection of SARS-CoV-2 RNA on currencies (both the prevalence study on circulating currency samples and persistance on spiked currencies).

| Target | Primer and probe sequences | Amplicon size |
| --- | --- | --- |
| ORF1b-nsp14 | Forward 5’-TGGGGYTTTACRGGTAACCT-3’ |  |
|  | Reverse 5’-AACRCGCTTAACAAAGCACTC-3’ |  |
|  | Probe 5’-FAM-TAGTTGTGATGCWATCATGACTAG-TAMRA-3’ |  |
| N | Forward 5’-TAATCAGACAAGGAACTGATTA-3’ |  |
|  | Reverse 5’-CGAAGGTGTGACTTCCATG-3’ |  |
|  | Probe 5’-ROX-GCAAATTGTGCAATTTGCGG-TAMRA-3’ |  |

Table A2: List of primers used for the PCR amplification and sequencing of different SARS-CoV-2 gene segments.

| Clade | Name | Primer Name | Sequence | Amplicon Size  (bp) | Tm (°C) |
| --- | --- | --- | --- | --- | --- |
| S | NS8_ 28144_ T to C | NS8_1_F | GTGGATGAGGCTGGTTCTAA | 217 | 54.6 |
|  |  | NS8_1_R | TGGGGTCCATTATCAGACAT |  | 53.6 |
| V | NS3_ 26144_G to T | NS3_3_F | CTGGTGTTGAACATGTTACCTT | 209 | 53.2 |
|  |  | NS3__3_R | CTCTTCCGAAACGAATGAGTA |  | 52 |
| G | Spike_ 23403_ A to G | Spike_1_F | CGTGATCCACAGACACTTGA | 228 | 54.6 |
|  |  | Spike_1_R | CCCTATTAAACAGCCTGCAC |  | 53.6 |
| GH | NS3_ 25563_G to T | NS3_ 2_F | CAAGGTGAAATCAAGGATGC | 207 | 51.8 |
|  |  | NS3_ 2_R | CAACAGCAAGTTGCAAACAA |  | 52.6 |
| GR | N protein 28882_G to A | N protein_1_F | AGGAACAACATTGCCAAAAG | 231 | 51.7 |
|  |  | N protein_1 R | TGTTGGCCTTTACCAGACAT |  | 54.2 |

Table A3: Conditions for the PCR amplification of different genes of SARS CoV 2

| Clade | PCR primer | Initial denaturation | Denaturation | Annealing | Extension | Number of cycles |
| --- | --- | --- | --- | --- | --- | --- |
| S | NS8_28144_T to C | 96 | 96 | 45 | 60 | 35 |
| V | NS3_ 26144_G to T |  |  | 45 |  |  |
| G | Spike_23403_AtoG |  |  | 52 |  |  |
| GH | NS3_ 25563_G to T |  |  | 45 |  |  |
| GR | N protein 28882_G to A |  |  | 51 |  |  |

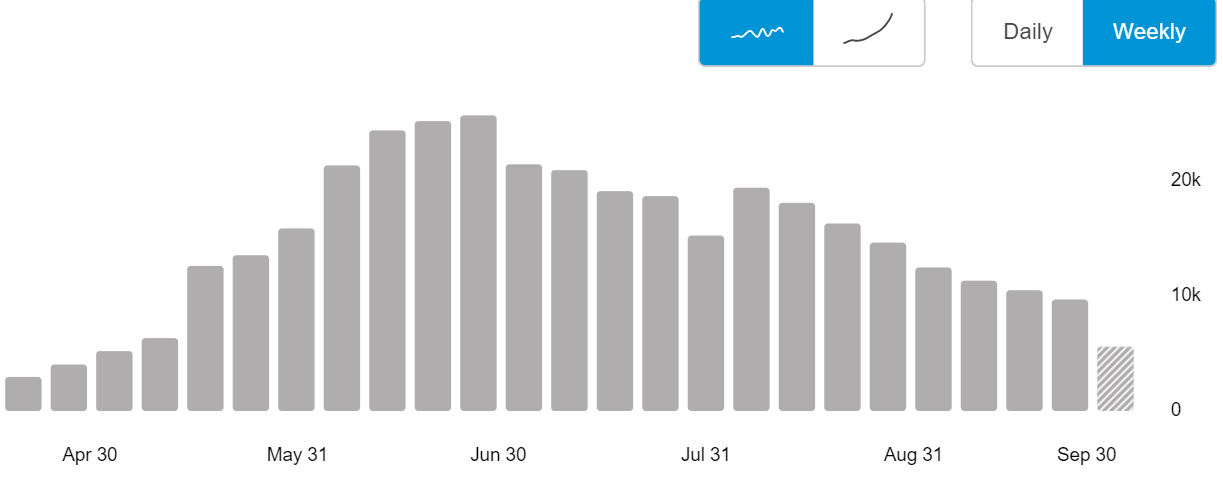

Figure A1: Weekly distribution of SARS CoV-2 confirmed case in Bangladesh from April to September 2020 (adopted from Corona dashboard, DGHS Bangladesh at https://dghs.gov.bd/)

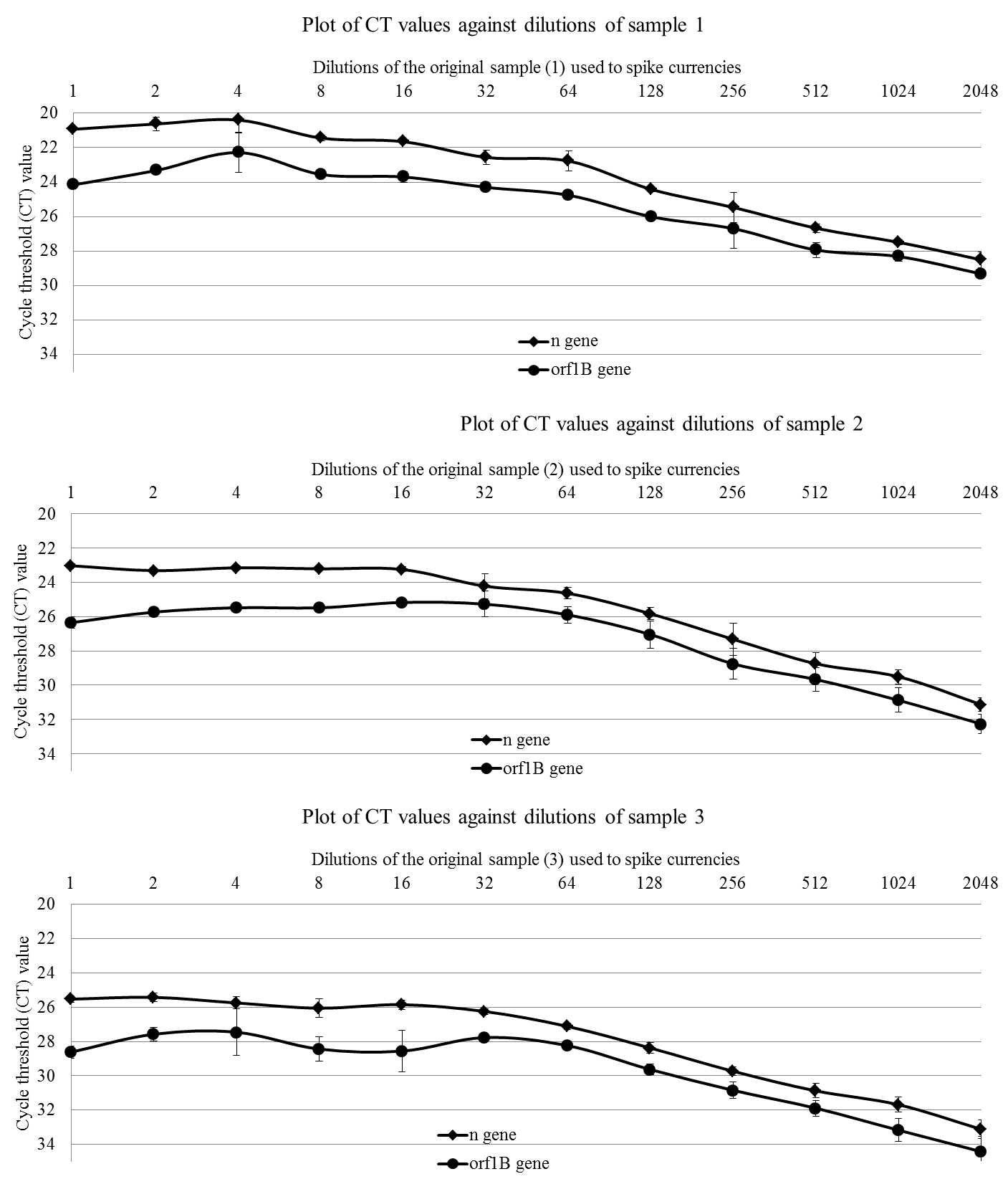

Figure A2: Standard curve plotted from the Cycle threshold values of the n- and orf1B –gene at 2-fold dilutions (detected by real time RT PCR assay) of the human nasopharyngeal samples used to spike the currency.
